# Populational analysis of the immunoglobulin G response to different COVID-19 vaccines in Brazil

**DOI:** 10.1101/2022.09.28.22280449

**Authors:** Nigella M. Paula, Marcelo S. Conzentino, Ana C.A. Gonçalves, Renata da Silva, Karin V. Weissheimer, Carlos H.S. Kluge, Paulo H.S.A. Marins, Haxley S.C. Camargo, Lucas R.P. Farias, Thamyres P. Sant’Ana, Letícia R. Vargas, Juliane D. Aldrighi, Ênio S. Lima, Guiomar T. Jacotenski, Fabio O. Pedrosa, Alan G. Gonçalves, Emerson Joucoski, Luciano F. Huergo

## Abstract

Vaccination is a strategy that confers protection against symptomatic infections and/or development of severe COVID-19. In Brazil, COVID-19 vaccination began in January 2021 and has been performed using vaccines from different manufactures including CoronaVac (Sinovac), ChAdOx1 (Oxford/AstraZeneca) and BNT162b2 (Pfizer/BioNTech). One of the main protective mechanisms triggered by vaccination involves the production of IgG antibodies reactive to the Spike antigen of SARS-CoV-2, the levels of which correlates with vaccine efficacy. Although phase III clinical studies confirmed the efficacy of the vaccines used in Brazil, there are just few studies comparing vaccine immunogenicity in a real-world scenario. This study aimed to depict the IgG response to natural infections and to vaccination using different types of vaccines at population scale in Matinhos, a city located in south of Brazil. Nucleocapsid seroconversion rates indicated that more than a quarter of the cohort has been subjected to natural infections by SARS-CoV-2 by the first trimester of 2022. Spike seroconversion rates achieved >95% by February 2022 and maintained stable as far as June 2022 confirming the effectiveness of the vaccination program. Immunogenicity concerning IgG reactive to Spike was higher using the BNT162b2 vaccine, followed by ChAdOx1 and CoronaVac. Natural infections boosted IgG levels reactive to Spike in those individuals that completed primary vaccination with ChAdOx1 and CoronaVac but not with BNT162b2. The levels of IgG reactive to Spike increased with the number of vaccine doses administered. The application of BNT162b2 as booster dose resulted in high levels of IgG reactive to Spike which were similar despite the type of the vaccine used during primary vaccination.

## INTRODUCTION

Since its isolation and description in the end of 2019, the severe acute respiratory syndrome coronavirus SARS-CoV-2 (Zhu *et al*. 2020), the causing agent of coronavirus disease COVID-19, has infected more than 600 million people and caused more than 6.5 deaths worldwide as by September 2022 https://coronavirus.jhu.edu/map.html. The high transmissibility of SARS-CoV-2 along with the rapid growing COVID-19 fatalities has led to massive global efforts to develop effective vaccines which would confer protection against symptomatic infections and/or development of severe COVID-19.

The main immunogenic components of SARS-CoV-2 are the Nucleocapsid (N) and Spike (S) proteins. Strong humoral IgG response against these antigens is detectable in COVID-19 convalescent cases (Ortega *et al*. 2021). The nucleocapsid protein is located within the viral particle in close contact with the viral RNA. On the other hand, the Spike protein is surface located and mediates the binding of SARS-CoV-2 to the angiotensin converting enzyme 2 receptor (ACE2) present on the host cell membrane. The Spike-ACE2 contact involves the Spike S1 reception-binding-domain (RBD) (Hu *et al*. 2021).

Due to the pivotal role of Spike in host-virus attachment, levels of antibodies reactive to Spike, particularly to S1 RBD, are linked with adaptative immunity protection (Planas *et al*. 2021). Therefore, most of the COVID-19 vaccines in use or under development were designed induce an immune response to the Spike antigen. Antibodies reactive to Spike may mediate protection not only by their ability to neutralize virus-host attachment but also through Fc-antibody-mediated process such as induction of phagocytosis, cytotoxicity and killer cell activation (Sadarangani, Marchant and Kollmann 2021).

Vaccines using different technologies became available to the general public in developed countries already in 2020. In Brazil, the public vaccination program begun at the end of January 2021. The vaccines available in Brazil were CoronaVac (Sinovac) which is based on inactive SARS-CoV-2; BNT162b2 (Pfizer/BioNTech) which is based on the novel mRNA technology; and ChAdOx1 (Oxford/AstraZeneca) and Ad26.COV2.S (Janssen) which are based on adenoviral vectors. The vaccines present only the full-length Spike protein to the immune system, the exception being CoronaVac which can elicit adaptative immune response to multiple SARS-CoV-2 antigens.

CoronaVac and ChAdOx1 were the first vaccines in use in Brazil becoming available in January 2021, followed by BNT162b2 in May 2021 and then by Ad26.COV2.S, which was introduced in June 2021 https://www.gov.br/saude/pt-br/coronavirus/vacinas/plano-nacional-de-operacionalizacao-da-vacina-contra-a-covid-19. The priority groups for vaccination were health-care workers, followed by groups with high-risk medical conditions and them by the general population from older to younger. The initial primary vaccination scheme was: CoronaVac two shots with 4-week interval; ChAdOx1 two shots with 12-week interval; BNT162b2 two shots with 12-week interval; Ad26.COV2.S was the only vaccine initially recommended as single dose. A booster 3^rd^ dose was recommended 150 days after primary vaccination.

Clinical studies involving the vaccines in use in Brazil reported efficacy against symptomatic infection after the 2^nd^ shot of 95% for BNT162b2 (Polack *et al*. 2020), 62-67% ChAdOx1 (Voysey *et al*. 2021), 50-84% CoronaVac (Tanriover *et al*. 2021) https://www.bbc.com/news/world-latin-america-55642648 (Cerqueira-Silva *et al*. 2022) and 70% Ad26.COV2.S (after 1 shot) (Sadoff *et al*. 2021). Despite these numbers, vaccine efficacy may be different in a real-world scenario as it may be affected by several factors to name a few: Challenging logistics of transportation and storage; Delays on second shot due to vaccine shortage and/or public hesitancy; Different genetic background and/or age groups of the population in relation to the cohort of phase III clinical trials; Surge of viral variants with significant immune scape. Therefore, depicting vaccine efficacy in real world scenario is an outstanding question.

One of the major mechanisms of protection elicited by vaccines involves the production of IgG antibodies reactive to Spike and to S1 RBD antigens, the levels of which are well correlated to each other as well as with virus neutralization activity. The levels of IgG reactive to Spike and S1 RBD are negatively correlated with the risk of symptomatic COVID-19 and hence, can be used to reliably predict the level of vaccine efficacy (Feng *et al*. 2021; Gilbert *et al*. 2022).

There are a few studies reporting that the COVID-19 vaccines in use in Brazil can elicit IgG reactive to the Spike antigen (Bochnia-Bueno *et al*. 2021; Medeiros-Ribeiro *et al*. 2021; Grenfell *et al*. 2022; Huergo *et al*. 2022). However, these studies were not designed to compare IgG levels obtained using different vaccines, they were limited to a small cohort and/or a specific group (i.e, heath care professionals) and/or did not provide information regarding the duration of the IgG response, the effectiveness of multiple vaccine doses/boosters nor to the effects of natural infections.

Here we investigated the IgG immunogenic response to Nucleocapsid, Spike and S1 RBD antigens of SARS-CoV-2 at a population scale in Matinhos, a touristic coastal city of the State of Paraná in the south of Brazil. Based on Nucleocapsid seroconversion rates we estimated that more than a quarter of the cohort has been subjected to natural infections by SARS-CoV-2 in the first trimester of 2022. Seroconversion rates for Spike reached >95% by February 2022 and maintained stable as far as June 2022. Spike and S1 RBD IgG levels were analyzed accordingly to the vaccine type, booster dose and natural infections. The results indicate a superior performance of BNT162b2, followed by ChAdOx1 and CoronaVac. Natural infections boosted IgG levels reactive to Spike in those individuals that completed primary vaccination with ChAdOx1 and CoronaVac but not with BNT162b2. The IgG levels reactive to Spike increased with the number of vaccine doses administered. We also noted that the levels of IgG reactive to Spike and S1 RBD were negatively correlated with age and interval after the second dose for the BNT162b2 vaccine.

## METHODS

### Study design and sampling

To understand the humoral response to SARS-CoV-2 in a real-world scenario no specific group/public were enrolled for this study. Any one older than 18 years was invited to participate. Participants could enroll at any time and multiple times for this study. The public was enrolled for the study by advertisement at university web site, radio and television. Sampling campaigns were performed weekly between January 2021 to June 2022 at the campus of Federal University of Paraná in the city of Matinhos.

Participants were directed to the study web site http://200.17.236.32/covid19/ where they could choose from any available date and time to come to the study center for the sampling campaigns. A questionnaire was presented online to collect personal information including age, gender and the city of residence; a self-declaration of any previous COVID-19 positive test and, if any, the date; a self-declaration of previous COVID-19 vaccine, the vaccine manufacture’s popular name (AstraZeneca, CoronaVac, Janssen or Pfizer) and the date of the first, second and third doses (if any). All information was stored in a MySQL database. Informed consent declaration was obtained from all participants. Ethics approval was obtained from the CEP/UFPR (n# 35872520.8.0000.0102) and CEP/HEG (n# 54095221.0.0000.0098).

### Serological analysis

Approximately 0.1 mL of blood was collected by capillary puncture. Samples were centrifuged (5,000 x g for 3 min) and 4 μL of serum was used to investigate IgG reactive against three different SARS-CoV-2 antigens Nucleocapsid (anti-N), Pre-fusion Spike (anti-S) and to Spike ΔS1 RBD (anti-RBD) (Uniprot MN908947, M153 to T589). All antigen sequences were based on the original Wuhan isolate. The details of antigen purification and bead preparation have been described previously (Conzentino *et al*. 2021a, 2021b, 2022; Huergo *et al*. 2021; Alvim *et al*. 2022).

All the SARS-CoV-2 antigens contained a 6x His-tagged and were coupled to Ni^2+^ magnetic beads. The presence of reactive IgG to anti-N and anti-S in the samples were investigated using an in-house ultra-fast high-throughput magnetic immunofluorescent assay operating at specificity >99.5%, sensitivity >95% as described previously (Conzentino *et al*. 2021b). The presence of high avidity IgG reactive to anti-S1 RBD were performed in chromogenic format under stringent conditions using urea 1 mol.l^-1^ in the wash buffers as described previously. Detection of IgG reactive to anti-RBD domain the specificity and sensitivity were >82% and 98% respectively (Conzentino *et al*. 2022).

Raw fluorescent reading (IgG reactive to anti-S and anti-N) or raw absorbance values (IgG reactive to anti-S1 RBD) in each sample were normalized using a reference serum and expressed as % of the reference. The positive cut off was set as 10%, 13% and 16% for protein Spike (S), Nucleocapsid (N) and Spike ΔS1 RBD (RBD), respectively.

### Statistical analysis

The data obtained from the questionnaire were exported to Microsoft Excel. Statistical analyses were performed using IBM SPSS version 28.0, NCSS 11, GraphPad Prism 8 and RStudio R 3.6.1 using the packages “FactoMineR” and “factoextra”. Multiple comparisons of IgG levels were performed in GraphPad Prism 8 using One-way ANOVA Tukey test, adjusted two-tailed p values are reported. Populational data confidence intervals were calculated considering positive prevalence of 10% for Nucleocapsid (N) and 50% for Spike (S) and Spike ΔS1RBD (RBD), the population size of 35,705 for the city of Matinhos was considered (https://www.ibge.gov.br/cidades-e-estados/pr/matinhos.html.

Seroprevalence results were compared with official numbers of reported COVID-19 cases and deaths occurring in the city of Matinhos and with the official numbers of applied vaccines in the state of Paraná. These data were available from the Paraná State Secretary of Health (https://www.saude.pr.gov.br/Pagina/Coronavirus-COVID-19). The estimated population of the Paraná state was 11,597,484 and the vaccine eligible population (>12 years old) in 2021 was estimated to be 8,883,672 based on the population pyramid predictions by the Brazilian Institute of Geography and Statistics IBGE (https://www.ibge.gov.br/apps/populacao/projecao/box_piramideplay.php?ag=41).

### Data availability statement

The data that supporting the findings of this study are available on request from the corresponding author (L.F.H).

## RESULTS

This study was set to depict the humoral immunoglobulin G response to different SARS-CoV-2 vaccines available in Brazil in a real-world scenario. Weekly sampling campaigns were opened to the public between January 2021 to June 2022 in the touristic city of Matinhos, in the coast region of the Paraná state, south of Brazil (Fig. S1). Any one older than 18 years old was invited to participate in this study. Participants could enroll at any time and multiple times.

A total of 2,376 samples were collected from 1,785 different individuals. All samples were analyzed for the presence of IgG reactive to nucleocapsid (N). Samples collected since July 2021 were also analyzed for the presence of IgG reactive to Spike (S) (n = 1,980). In addition, 1,225 samples were further evaluated for the presence of high avidity antibodies reactive to S1 RBD. In total, 5,581 analyses were performed. Although the study site was in the city of Matinhos, visitors and workers residing in other cities were also enrolled. The cohort was composed of residents of Matinhos 60%, Curitiba 26% (capital of the Paraná state), Paranaguá 6.4% (main city of the Paraná coast), Guaratuba 3.5% and from other cities 2.8% (Figure S1). The age of the participants had a mean of 40 years (SD 13; min. 18 max. 86 years old), with a 65% predominance of women.

### Seroconversion rates for Nucleocapsid, Spike and S1 RBD

To understand the effectiveness of the vaccination program in Brazil it is important to estimate the fraction of the population that had experienced natural infection. These numbers can be determined based on the fraction of those individuals with a positive IgG test for the N antigen after excluding individuals vaccinated with CoronaVac, the only vaccine applied in Brazil that can elicit anti-N antibodies. In our previous study, serological analysis indicated that 16.7% (12.6 – 20.7, 95% CI) of the population had experienced COVID-19 by the end of 2021 (Huergo *et al*. 2022). This number raised sharply in 2022 after the introduction of the SARS-CoV-2 omicron variant in the region (Adamoski *et al*. 2022), reaching 26.1% (23.3 – 28.5, 95% CI) in the first trimester of 2022. The number remained stable in the second trimester of 2022, 25.6% (21.4 – 29.8, 95% CI) (Figure S2). The data indicate that more than a quarter of the non-vaccinated cohort had experienced SARS-CoV-2 infection by March 2022. It is worth mentioning that the real number of cases should be higher as IgG anti-N sero-revertes had been detected in this cohort during our previous study (Huergo *et al*. 2022).

We showed in our previous study that Spike seroconversion in 2021 followed the trend of the population fraction that had taken the second dose (Huergo *et al*. 2022). The numbers in 2022 confirmed this trend. By February 2022 official numbers indicated that 96.4% of the eligible population had completed the primary vaccination (2^nd^ dose). Accordingly, 95.6% (88.3 – 100%, 95% CI) of Spike seroconversion was detected. Spike seroconversion remained above 95% between February and June 2022 (Fig. S2B). Seroconversion for high avidity IgG reactive to SARS-CoV-2 S1 RBD antigen was 53.8% (48.6 – 58.9, 95% CI) in November 2021 reaching 70.4% (62.2 – 78.6, 95% CI) in February 2022. The numbers remained stable between 65-70 % from February to June 2022 (Fig. S2B).

The data described above support the effectiveness of the Brazilian vaccination program concerning Spike seroconversion. However, it is important to stress that a significant fraction of Spike seroconverts can result from natural infections, estimated to have occurred in more than a quarter of the cohort at first trimester 2022. In addition, different priority groups received vaccines from different manufacturers as they were made available in Brazil, hence, it is important to understand the rate of seroconversion and the levels of IgG considering different vaccines and/or priority groups.

### IgG reactive to Spike and S1 RBD upon completion of primary vaccination

To depict the IgG response raised after completion of the primary vaccination (2^nd^ dose) using vaccines from different manufactures the cohort was stratified following the information provided by the participants. From this point on, unless stated otherwise, the following filters were applied to the cohort. Participants who self-declared previous COVID-19 positive test were excluded to minimize inputs resulting from natural infections. Only participants who declared to have taken the second dose of the vaccine in the time frame between 10 and 240 days before sampling were considered. Those that took booster doses were excluded. After applying the above-mentioned filters, the remaining cohort consisted of 804 samples from 716 individuals distributed accordingly to the vaccine type as ChAdOx1 (AstraZeneca) 45.4%, BNT16b2 (Pfizer) 32.7%, CoronaVac (Sinovac) 21.6% and Ad26.COV2.S (Janssen) 0.2%. Given the low representative of Ad26.COV2. S, this vaccine type was excluded from the comparative analyses.

The IgG seroconversion rates in participants vaccinated with BNT16b2 were 99% and 77% for Spike and S1 RBD, respectively. ChAdOx1 positive rates were 81% for Spike and 36% for S1 RBD. Volunteers vaccinated with CoronaVac were 52% and 31% positive for Spike and S1 RBD, respectively. This trend in seroconversion ratio was evident when the signal of IgG reactive to Spike and S1 RBD were plotted accordingly to the vaccine type. The IgG levels were higher for BNT16b2, followed by ChAdOx1 and CoronaVac. The differences were significant (p<0.001) in all comparisons except for ChAdOx1 vs CoronaVac in the case of IgG reactive to S1 RBD (p= 0.46) (Fig. 1).

**Fig. 1:**
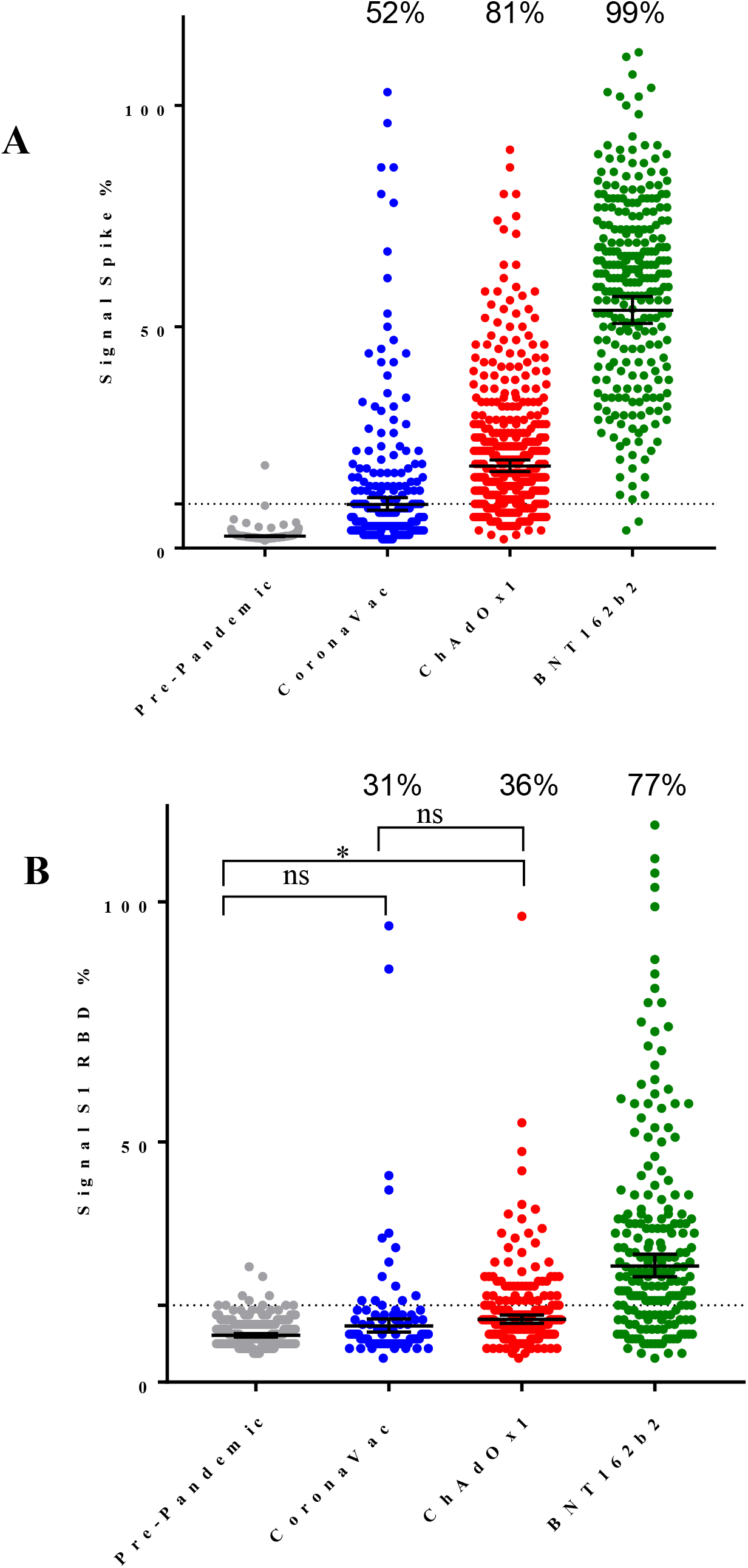
The IgG response to Spike and S1 RBD accordingly to the vaccine type. IgG levels reactive tp Spike (A) and S1 RBD (B) in participants negative for COVID-19 which completed primary vaccination (2^nd^ dose) between 10 to 240 days. Bars represent geometric mean with 95% CI, the dashed line indicates the seropositive cutoff. Pre-pandemic samples were plotted as naïve controls. One-Way ANOVA analysis were performed, all comparisons were significant at p<0.0001 except when indicated in the graphic (* p<0.05), (ns, not significant). Numbers indicate seroconvertion rates

It is important to note that the average IgG levels for Spike and S1 RBD were significantly higher (p<0.001) after completion of primary vaccination using vaccines from different manufactures, when compared to pre-pandemic samples (Fig. 1). The only exception was CoronaVac were IgG reactive to S1 RBD was not significantly different from pre-pandemic samples (Fig 1). These data confirm that all vaccines in use in Brazil were effective in activating the IgG response to the Spike antigen. The levels of IgG reactive to Spike and RBD were not influenced by gender in different vaccine types (Fig. S3).

### IgG response according to age and interval after the second dose

Important questions regarding humoral response obtained with different types of vaccines are: How does the IgG response perform at different age groups? Does the IgG response wane over time? To obtain insights into these questions Pearson correlation analysis were performed by plotting the Spike or RBD IgG signal versus age or versus time interval after completion of primary vaccination (2^nd^ dose). For the CoronaVac and ChAdOx1 vaccines there were no significant correlation between IgG levels reactive to Spike or RBD with age or time interval (Fig. 2 and S4). On the other hand, for the BNT16b2 vaccine, significant (p<0.0001) weak to moderate negative correlation were observed in these comparisons (Fig. 2 and S4). Thus, even though BNT16b2 performed better than CoronaVac and ChAdOx1 to induce humoral IgG response to Spike and RBD, the ability of BNT16b2 to sustain such high response was negatively influenced by age and the time after the second dose.

**Fig. 2:**
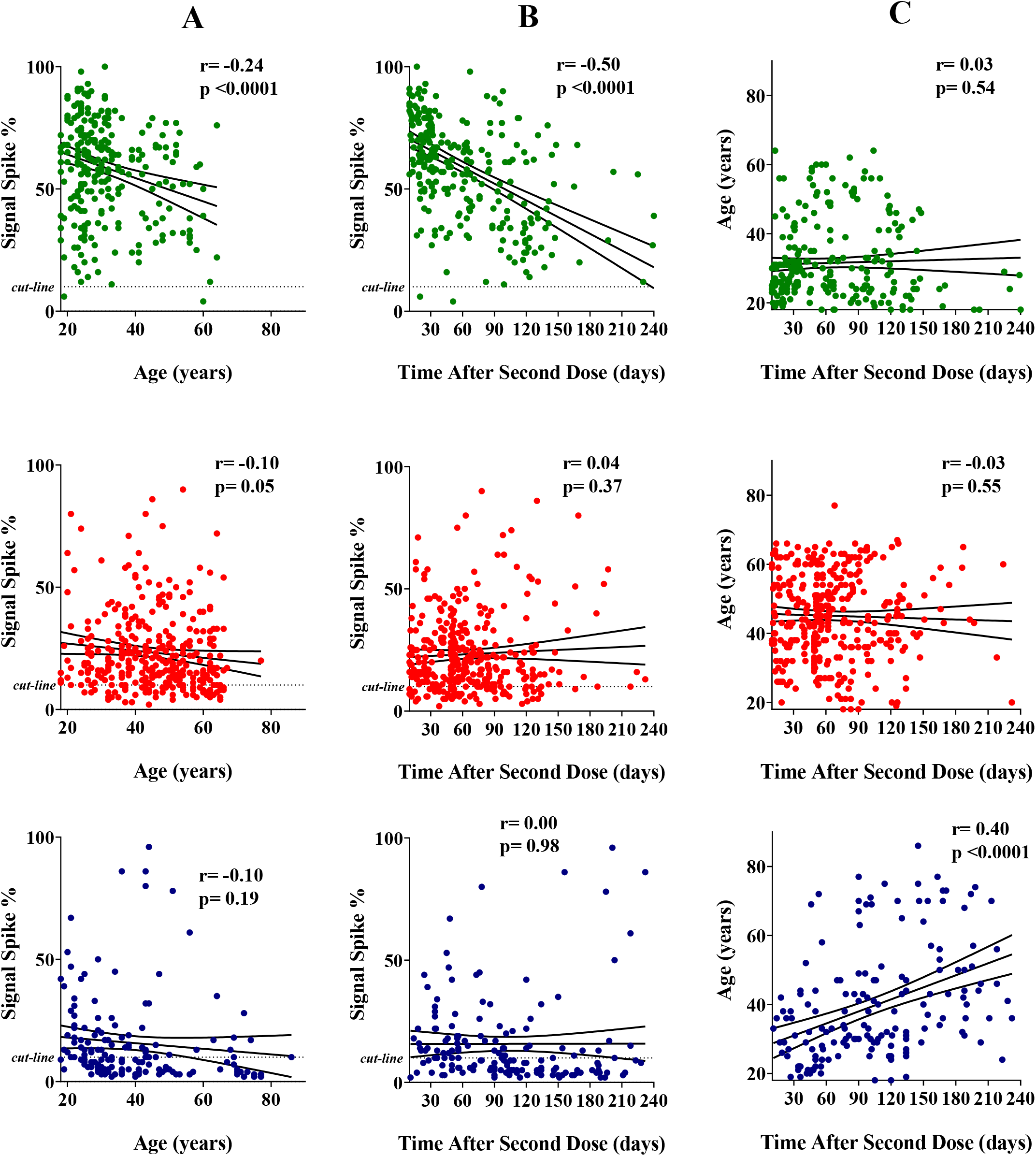
Correlation between the levels of IgG reactive to Spike and the age or time after the second vaccine dose. The IgG levels were determined in participants negative for COVID-19 which completed primary vaccination (2^nd^ dose) between 10 to 240 days. (A) Correlation between IgG levels and age. (B) Correlation between IgG levels and time after the second dose. (C) Correlation between Age and time after the second dose. The linear regression with 95% CI and Pearson’s correlation coefficient is indicated. Color code, green BNT162b2, red ChAdOx1, blue CoronaVac.

The distribution of age and interval were not even within the x axis among different vaccine types (Fig. 2). Furthermore, in the CoronaVac subgroup, there was a significant positive correlation between age and time interval after the second dose (Fig. 2). This is explained by the fact that CoronaVac was the first vaccine introduced in Brazil and mainly applied to elderly. To compare vaccine immunogenicity performance without age and/or interval bias the cohort was further filtered to those individuals of 18-40 years that received the second dose between 10-90 days before sampling. When the levels of IgG reactive to Spike and S1 RBD in this subgroup were plotted accordingly to the vaccine type the same trend and statistics presented in Fig. 1 was detected (Fig. S5). This data confirms that that BNT16b2 was more effective in the immediate activation of the humoral response followed by ChAdOx1 and CoronaVac.

### Comparison of primary vaccination and natural infection accordingly to vaccine type

Individuals that completed the primary vaccination (2^nd^ dose) were distributed accordingly to the vaccine type and separated in infected and non-infected subgroups (Fig. 3A). For this analysis IgG reactive to RBD was not considered due to low number of samples in some subgroups. Participants who declared a previous positive diagnostic for COVID-19 were considered as the infected group despite infections occurred before or after 1^st^ or 2^nd^ dose of the vaccine.

**Fig. 3.**
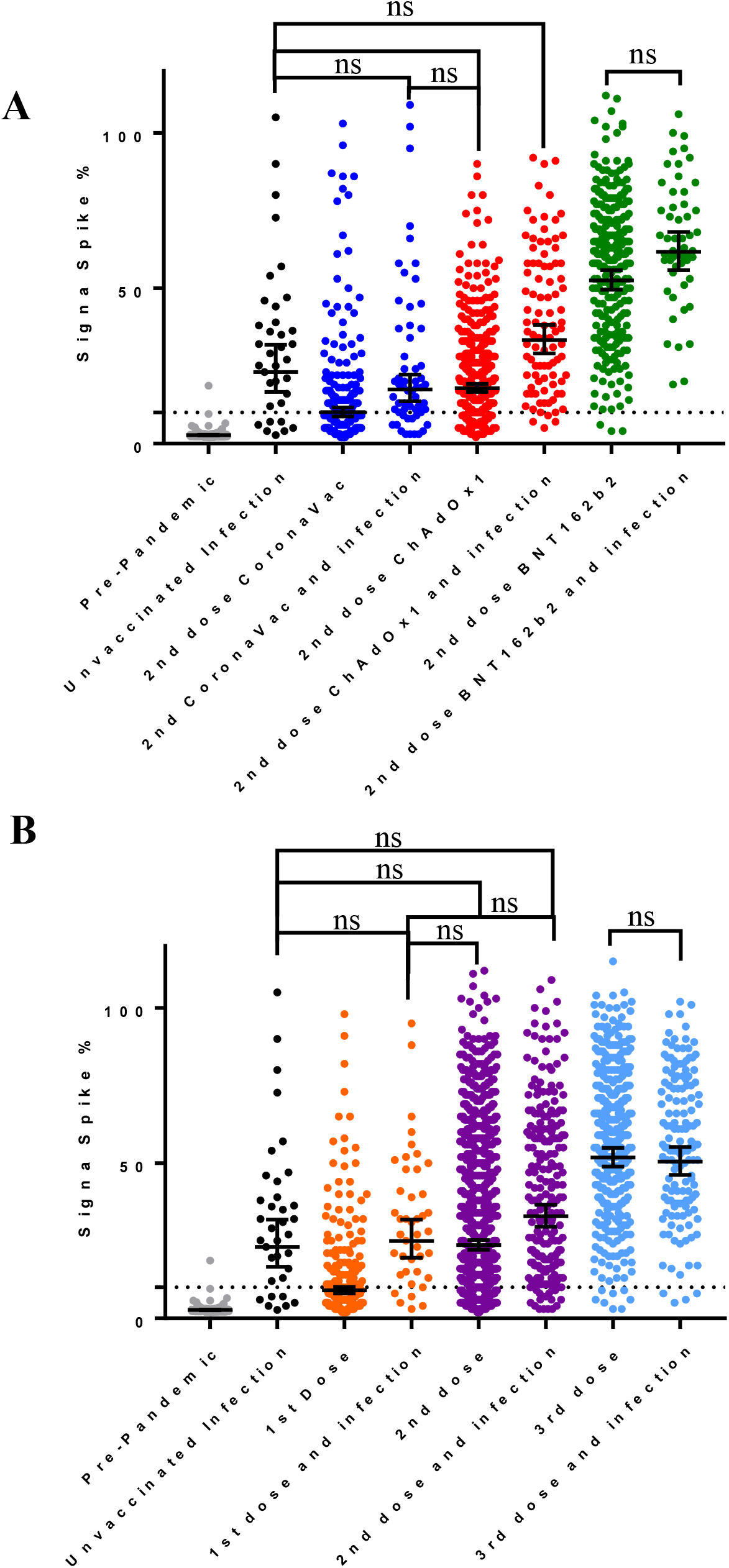
The levels of IgG reactive to Spike in response to infection and vaccination. A) Participants that completed the primary vaccination (2^nd^ dose) were grouped accordingly to the vaccine type. Those participants self declaring a previous positive COVID-19 test are indicated as infection cases. B) Participants were grouped accordingly to the number of vaccine doses. The dashed line indicates the seropositive assay cutoff. Pre-pandemic samples were plotted as naïve controls. Bars represent geometric mean and 95% CI. One-Way ANOVA analysis were performed comparisons were all significant (p<0.02) unless indicated as not significant (ns).

Natural infections increased IgG levels reactive to Spike in the CoronaVac (p=0.017) and ChAdOx1 (p<0.0001) subgroups but not in those individuals vaccinated with BNT16b2 (p=0.17). Comparing IgG levels between infected non-vaccinated vs non-infected vaccinated individuals indicated that natural infections resulted in higher, equal, and lower IgG levels than those obtained upon completion of primary vaccination with CoronaVac (p=0.001), ChAdOx1(p=0.06) and BNT16b2 (p<0.0001), respectively.

### IgG response to infection vs multiple dose vaccination

The complete dataset was used to evaluate the effect of natural infections and multiple vaccine doses on the levels of reactive IgG for Spike. Again, the IgG reactive to RBD was not considered due to low sample size in some subgroups. Participants who declared a previous positive diagnostic for COVID-19 were considered as the infected group.

Participants were grouped accordingly to the number of vaccine doses received with all types of vaccines combined. In the non-infected group, participants presented increased levels of IgG reactive to Spike accordingly the number of vaccine doses (Fig. 3B). The pairwise comparison with increasing dose number (i.e, 0 vs 1; 1 vs 2, etc) were all statistically significant (p<0.0001). Within the infected group, increased IgG levels were significant (p<0.0001) only for the 2^nd^ dose vs booster (3^rd^ dose) comparison (Fig. 3B). Pairwise comparison, considering the same number of doses, between infected and non-infected individuals showed that the natural infection increased IgG levels significantly (p<0.0001) for pre-pandemic unvaccinated, 1^st^ and 2^nd^ doses. However, upon administration of the 3^rd^ dose (booster), IgG levels became similar (p>0.99) between infected and non-infected groups.

### Effect of booster (3^rd^ dose) accordingly to the vaccine type

We evaluate the effect of the booster dose according to the type of vaccine applied. Participants who declared to have taken the 3^rd^ dose of the vaccine in the time frame between 10 and 240 days before sampling were considered. Given that after the 2^nd^ and 3^rd^ vaccine doses natural infections are more likely to be asymptomatic, participants were included despite the declaration of previous infection in these analyses. For most participants (90%), the 3^rd^ dose was BNT16b2 including homologous and heterologous combinations of primary vaccination. A small number of participants (8.7%) had a 3^rd^ dose of ChAdOx1 combined with homologous ChAdOx1 primary vaccination. Other combinations were present in small numbers and thus were not considered in the comparisons.

The 3^rd^ dose heterologous vaccination regimes ChAdOx1 + BNT16b2 and CoronaVac + BNT16b2 were very effective in augmenting the IgG levels and Spike seroconversion rates in comparison to primary vaccination (p<0.0001). In fact, antibody levels achieved were similar to those obtained with the homologous BNT16b2 + BNT16b2 regime (Fig. 4).

**Figure 4:**
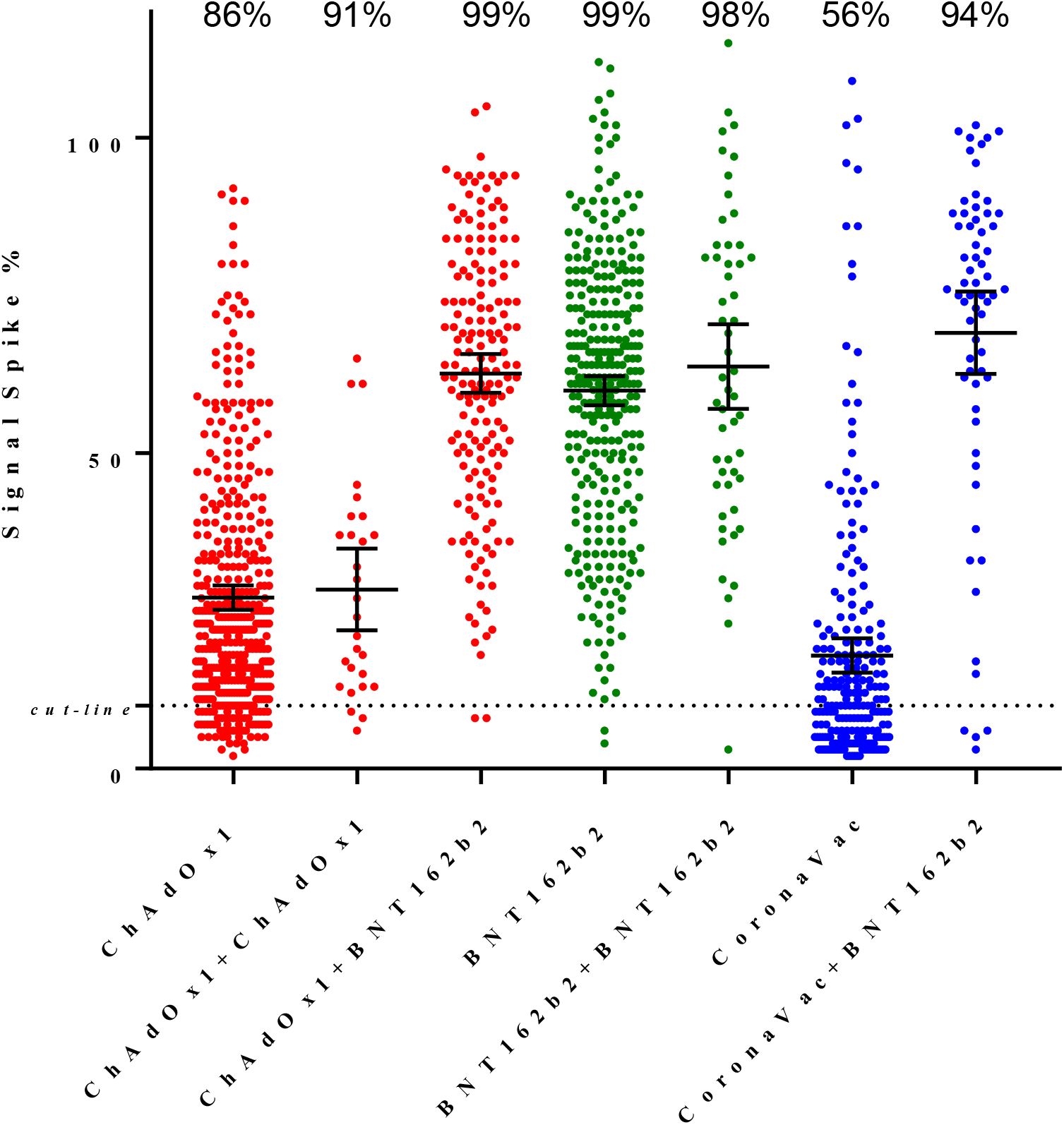
Comparison of IgG levels reactive Spike after completion of primary vaccination and after the first booster (3^rd^ dose) accordingly to the vaccine type. IgG levels reactive for Spike in all participants (including those negative and positive for natural infections) which completed primary vaccination (2^nd^ dose) between 10 to 240 days are indicated accordingly to the vaccine type. The IgG levels on participants that toke a booster 3^rd^ dose are indicated with + followed by the vaccine type of the booster. Only participants that took the booster dose between 10 to 240 days were considered. The dashed line indicates the seropositive assay cutoff. The geometric mean and 95% CI is represented by the bars. Number indicate the seroconversion rates.

The 3^rd^ dose homologous regimes ChAdOx1 + ChAdOx1 and BNT16b2 + BNT16b2 were able to sustain, but not increase, the IgG levels in comparison to primary vaccination (Fig. 4). This data indicates that the homologous regime BNT16b2 + BNT16b2 was superior to ChAdOx1 + ChAdOx1 concerning levels of IgG reactive to Spike (p<0.0001) (Fig. 4).

### Principal component analysis

The full data set was subjected to principal component analysis (PCA) to identify the major variables. The dimension 1 (x axis), which explained 27% of the variation, had as main vectors the variables vaccination, number of doses, time after doses and IgG signal to Spike and RBD (Fig. S6). All these variables were pointing to the positive side of the x axis suggesting correlation among as expected and confirming the trend reported in previous analyses. The dimension 2 (y axis) explained 11% of the variation and had as the main vectors the variables a diagnostic for COVID-19, the time after the positive diagnostic and the signal of IgG reactive to Nucleocapsid. The vectors of these two variables were nearly superimposed (Fig. S6) indicating excellent correlation. The PCA analysis confirmed that gender and city of residence resulted in negligible vectors which did not influence dataset variability whereas age had a minor contribution.

## DISCUSSION

Global efforts for COVID-19 vaccine development resulted in an extraordinary number of vaccines candidates developed. Many countries, including Brazil, adopted vaccines based on different technologies from different manufacturers. To understand how these different vaccines perform in a real-world scenario is an outstanding question. It is assumed that the levels of IgG reactive to Spike and S1 RBD can be used to reliably predict vaccine efficacy (Feng *et al*. 2021; Gilbert *et al*. 2022).

Previous studies dealing with COVID-19 vaccine immunogenicity were focused on a particular cohort, type of vaccine and/or pre-defined dose intervals. The project that gave rise to the present work was primarily defined as an action of public health, with the objective to awake the interest of citizens on vaccination. As such, our cohort was based on a broader unprompted range of participants, of which, variables originated from real-world scenario, where different vaccines were used for different priority groups in a context concurring with natural infections and vaccine hesitancy.

We first analyzed the evolution of SARS-CoV-2 infections and vaccination by using seroconversion to Nucleocapsid and Spike as a proxy, respectively. The data allowed to estimate that more than a quarter of the population had been infected by SARS-CoV-2 in the first trimester of 2022 (Fig S1A). The effectiveness of the vaccination program concerning Spike immunogenicity is clear as Spike seroconversion achieved >95% in February 2022 remaining at this level up to the end of this study by June 2022 (Fig S1B). Despite the high Spike seroconversion rates, a sharp increase in COVID-19 cases were detected between January-March 2022 (Fig. S1A and C). These cases are attributed to the omicron variant which was predominant in the region at this time (Adamoski *et al*. 2022) and has the documented ability to evade neutralizing antibodies raised by vaccines and/or prior infections (Planas *et al*. 2021; Carreño *et al*. 2022).

Despite the high number of COVID-19 cases between January-March 2022, it did not reflect in increased number of deaths (Fig. S1C). The lower fatality rate can be partially attributed to the fact that the omicron variant causes less severe infections due to reduced viral replication in the lungs (Hui *et al*. 2022). Furthermore, the presence of preexisting Spike binding antibodies in the population (Fig. S1B), despite the low neutralizing activity against omicron (Grenfell *et al*. 2022), are likely to be a key factor to reduce case fatality by reducing viral replication through Fc-mediated processes (Sadarangani, Marchant and Kollmann 2021).

The immunogenicity of the different vaccines upon completion of primary vaccination was compared in the population. This analysis revealed that both seroconversion rates and the levels of IgG reactive to Spike and S1 RBD were higher on those participants vaccinated with BNT16b2, followed by ChAdOx1 and CoronaVac, in agreement with other studies (Barin *et al*. 2022; Romero-Pinedo *et al*. 2022).

Quite remarkably, the seroconversion rates for Spike binding antibodies determined in our study (Fig. 1A) is in excellent agreement with vaccine efficacy numbers reported previously. Vaccine efficacies vs seroconversion rates concerning primary vaccination were: BNT16b2 95% (Polack *et al*. 2020) vs 99%; ChAdOx1 71% (study in Brazil) (Voysey *et al*. 2021) vs 81%; Coronavac 50-55% (study in Brazil) https://www.bbc.com/news/world-latin-america-55642648 (Cerqueira-Silva *et al*. 2022) vs 52%. This was also observed after booster doses. For instance, combining CoronaVac + BNT16b2 resulted in vaccine efficacy against infection of 93% (study in Brazil) (Cerqueira-Silva *et al*. 2022) whereas Spike seroconversion was 94% (Fig. 4).

The above mentioned data reinforces the idea that vaccine efficacy correlates with the levels of IgG reactive to Spike (Feng *et al*. 2021; Gilbert *et al*. 2022), furthermore it suggests that vaccine efficacy numbers reported in clinical studies can be extrapolated to the real-word scenario in Brazil. Readers should be cautioned, however, that a positive Spike IgG test may not represent an immune passport to symptomatic infections, especially in the light of emerging SARS-CoV-2 variants which exhibit exceptional ability to evade pre-existing antibodies. Furthermore, studies indicate that vaccine effectiveness and IgG response wane over time and are negatively influenced by age (Choudhary *et al*. 2021; Levin *et al*. 2021; Andrews *et al*. 2022).

In the current study we noted a significant negative correlation between age or interval after the second dose with the levels of IgG reactive to Spike and RBD for BNT16b2. Strikingly, such effect could not be detected in participants that were vaccinated with ChAdOx1 and CoronaVac (Fig. 2 and S4). It is not clear, however, if the lack of negative correlation in the case of ChAdOx1 and CoronaVac are linked to the fact that these vaccines elicited a much lower initial response than BNT16b2. Other studies in Brazil identified that both vaccine efficacy and anti-Spike IgG levels decreased >180 days after primary vaccination with CoronaVac (Cerqueira-Silva *et al*. 2022; Grenfell *et al*. 2022). It is worth mentioning that our study is limited by the fact the longitudinal data was determined among different participants which were predominantly younger than 60 years. Furthermore, there were a limited number of samples converting >180 days after vaccination in our study.

Data reported in this study confirms previous findings that natural infections act as additional booster to raise the levels of IgG reactive to Spike (Grenfell *et al*. 2022). The raise in IgG levels was clearly observed on those individuals that completed primary vaccination with CoronaVac and ChAdOx1 but not with BNT16b2, for which, primary vaccination alone elicited a high humoral response (Fig. 3).

Application of a vaccine 3^rd^ dose raised the levels of IgG reactive to Spike and seroconversion rates. The levels of IgG after the 3^rd^ vaccine dose were at such high scale that no additional booster effect could be visible caused by a natural infection (Fig. 3). The combination of different types of vaccine during primary vaccination and booster doses indicated that the use of BNT16b2 as booster resulted in increased seroconversion rates and IgG levels when primary vaccination was completed with CoronaVac or ChAdOx1, which is in agreement with previous finding in Brazil (Costa Clemens *et al*. 2022). Such raise on IgG was not detected when ChAdOx1 was used as booster in homologous vaccination regime (Fig. 4).

One important limitation of this study was the fact that participants were not randomly selected in the population, they spontaneously volunteered and came to the study site. Hence, populational bias may imply. Other aspects to consider is that time comparative analysis such was waning IgG levels over time (Fig. 2) represent the general population and not a prospective cohort.

In conclusion, assuming the premise that the anti-Spike IgG levels act as a proxy for vaccine efficacy and protection against COVID-19, our time-series Spike seroconversion data (Fig. S1B) confirmed the effectiveness of the Brazilian vaccination program was key to control the SARS-CoV-2 pandemy. The adhesion of the population to both primary vaccination campaign and booster shots were key to maintain high humoral protection. Of the vaccines used in Brazil, BNT16b2, which is based on the novel mRNA technology, performed better to elicit anti-Spike IgG after primary vaccination and as a booster dose and thus should be recommended as booster whenever available.

## Data Availability

All data produced in the present study are available upon reasonable request to the authors

## ACKNOWLEDGMENT

We thank all the volunteers that contributed samples to this work. We acknowledgment the financial support of Alexander von Humboldt foundation, UFPR, CNPq, CAPES and Fundação Araucária. We acknowledge Profa Leda Castilhos, Laboratório de Engenharia de Cultivos Celulares (LECC) Coppe, UFRJ Rio de Janeiro, RJ, Brazil for providing the Spike protein used in this study.

## AUTHOR’S CONTRIBUTION

M.C, A.G., R.S., K.W, L.F., T.S., L.V., J.A., E.L., G.J, collected the data and performed serological analyses. C.K., P.A., H.C., database creation and maintenance: E.J. statistical analysis. N.P. collected and analyzed the data. L.H, conceptualisation, formal analysis, funding acquisition, collected and analysed the data. L.H and N.P wrote the paper with inputs from F.P., A.G. and N.P.

---

**Figure S1:**
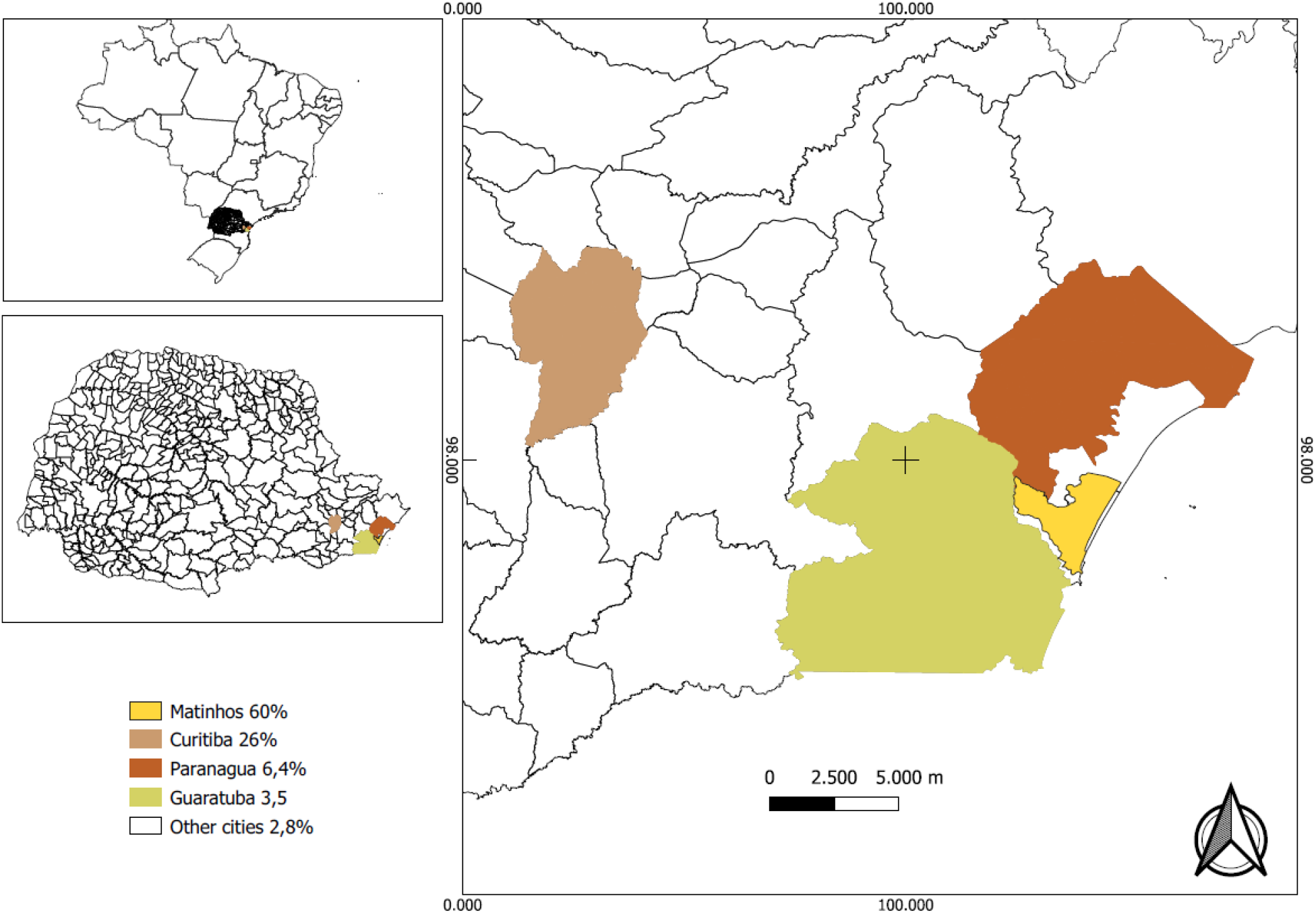
Location of the study site and city of residence of the participants.

**Fig. S2:**
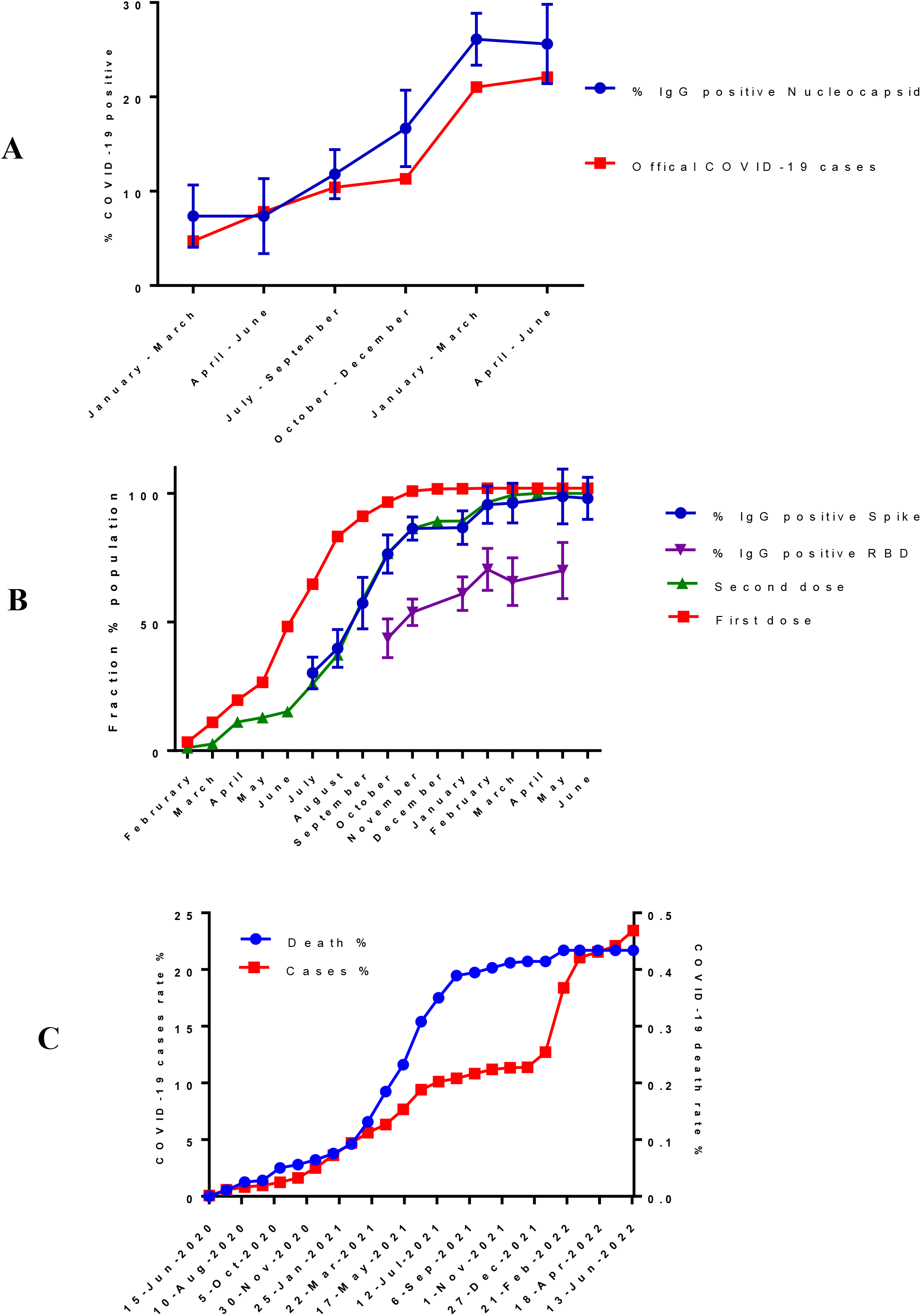
Evolution of COVID-19 infection, vaccination and humoral response. (A) Evolution of COVID-19 during each quarter from January 2021 to June 2022. The percentage of samples IgG positive to the nucleocapsid antigen is represented by the blue line (error bars confidence levels at a 95% CI). The red line indicates the percentage of official cases of COVID-19 reported by health authorities in the city of Matinhos (cumulative cases in the middle of each quarter are reported as a percentage of the population). (B) The fraction of the population (>12 years) vaccinated with first dose (red), with the second dose (green) in the State of Paraná. The IgG seroconversion rate to Spike (blue) and RBD (purple). Error bars indicate confidence levels at a 95% CI. (C) Monthly evolution of official cases of COVID-19 (red) and death (blue) in the city of Matinhos.

**Fig. S3:**
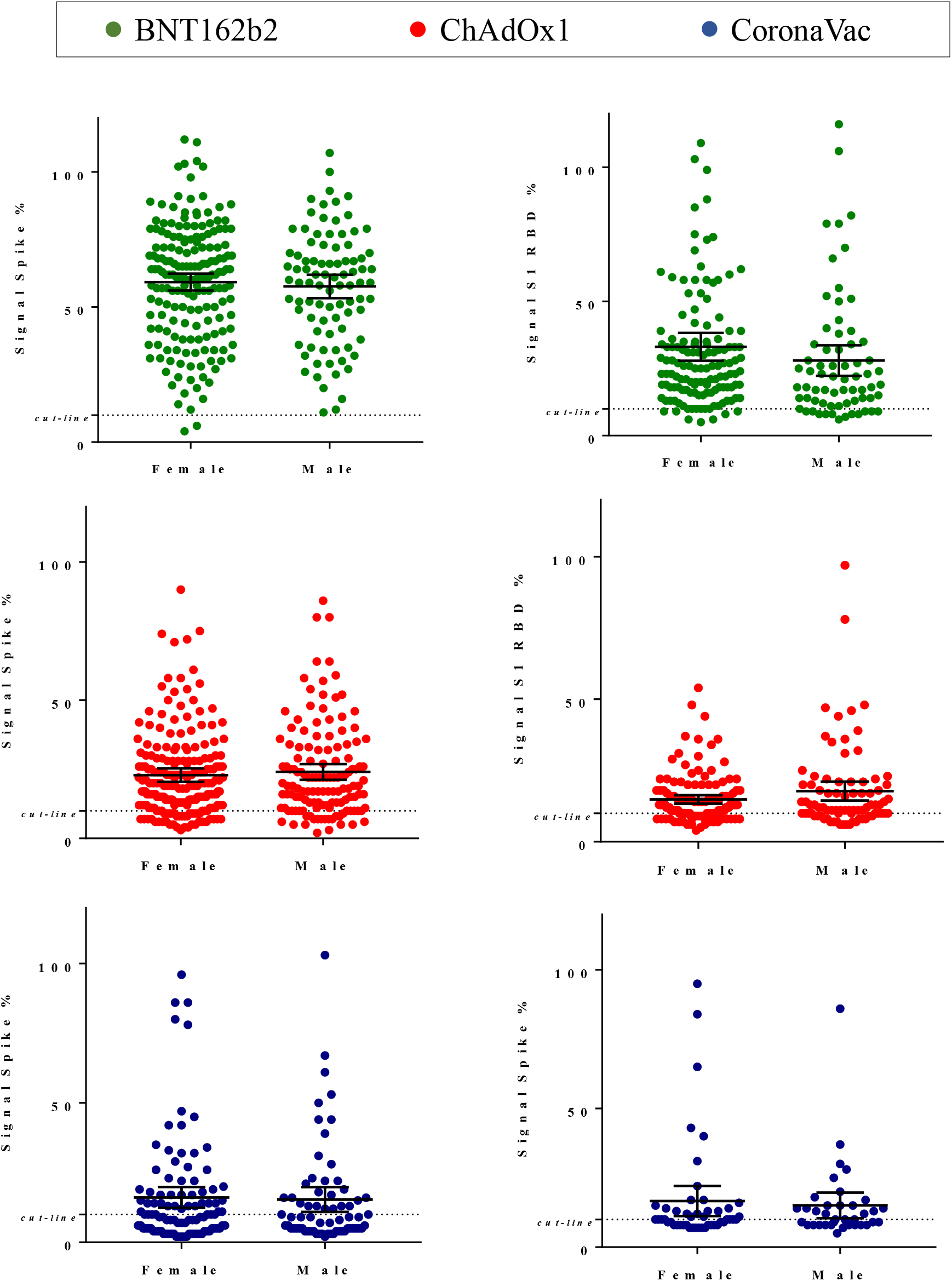
IgG response to Spike and RBD according to gender. IgG levels were determined in participants negative for COVID-19 which completed primary vaccination (2^nd^ dose) between 10 to 240 days. The geometric mean is indicated with error bars reporting 95% CI. Gender was not statistically significant in all comparisons (p>0.05) in paired t-test.

**Fig. S4:**
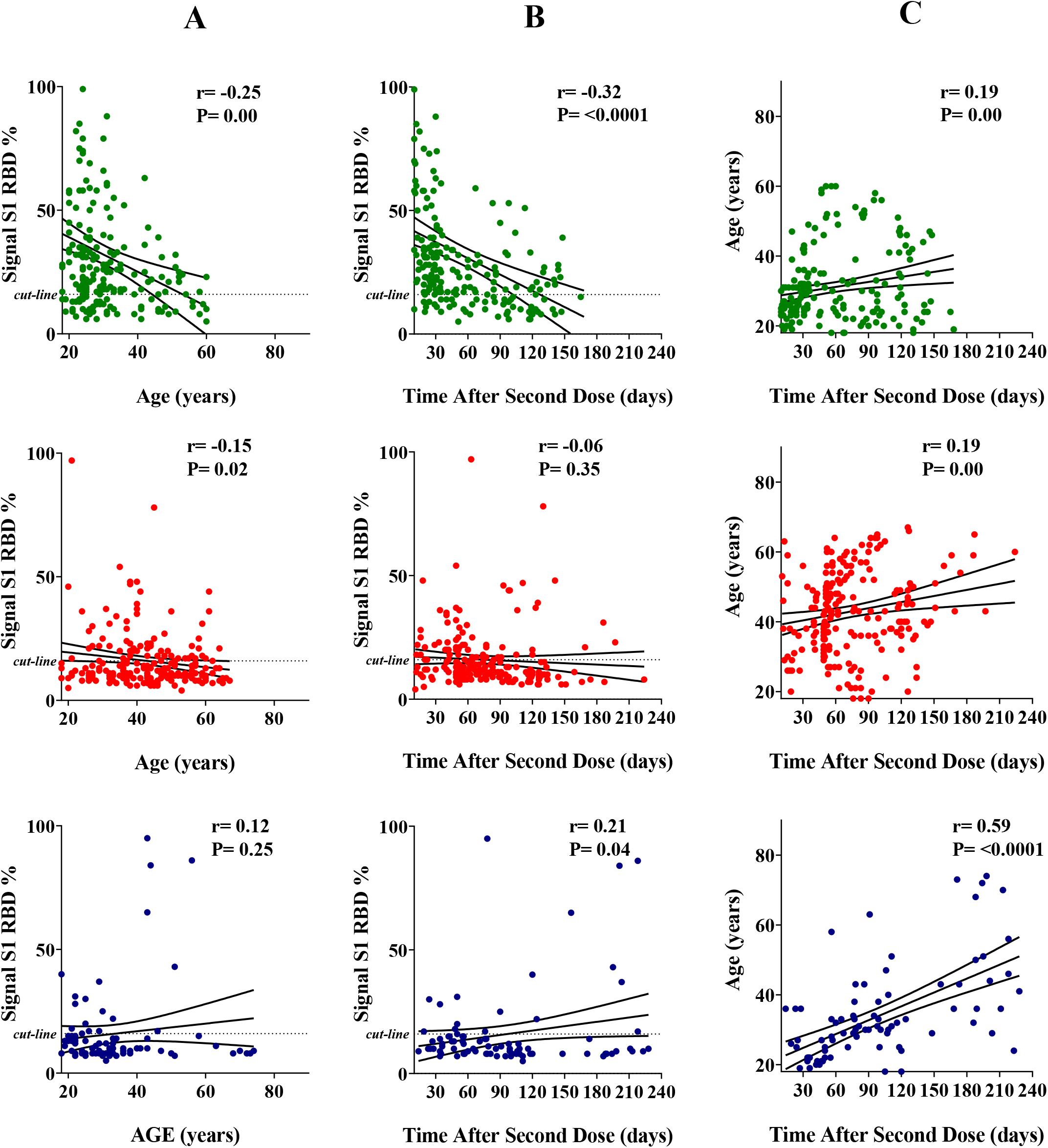
Correlation between the levels of IgG reactive to S1 RBD and the age or time after the second vaccine dose. The IgG levels were determined in participants negative for COVID-19 which completed primary vaccination (2^nd^ dose) between 10 to 240 days. (A) Correlation between IgG levels and age. (B) Correlation between IgG levels and time after the second dose. (C) Correlation between Age and time after the second dose. The linear regression with 95% CI and Pearson’s correlation coefficient is indicated. Color code, green BNT162b2, red ChAdOx1, blue CoronaVac.

**Figure S5:**
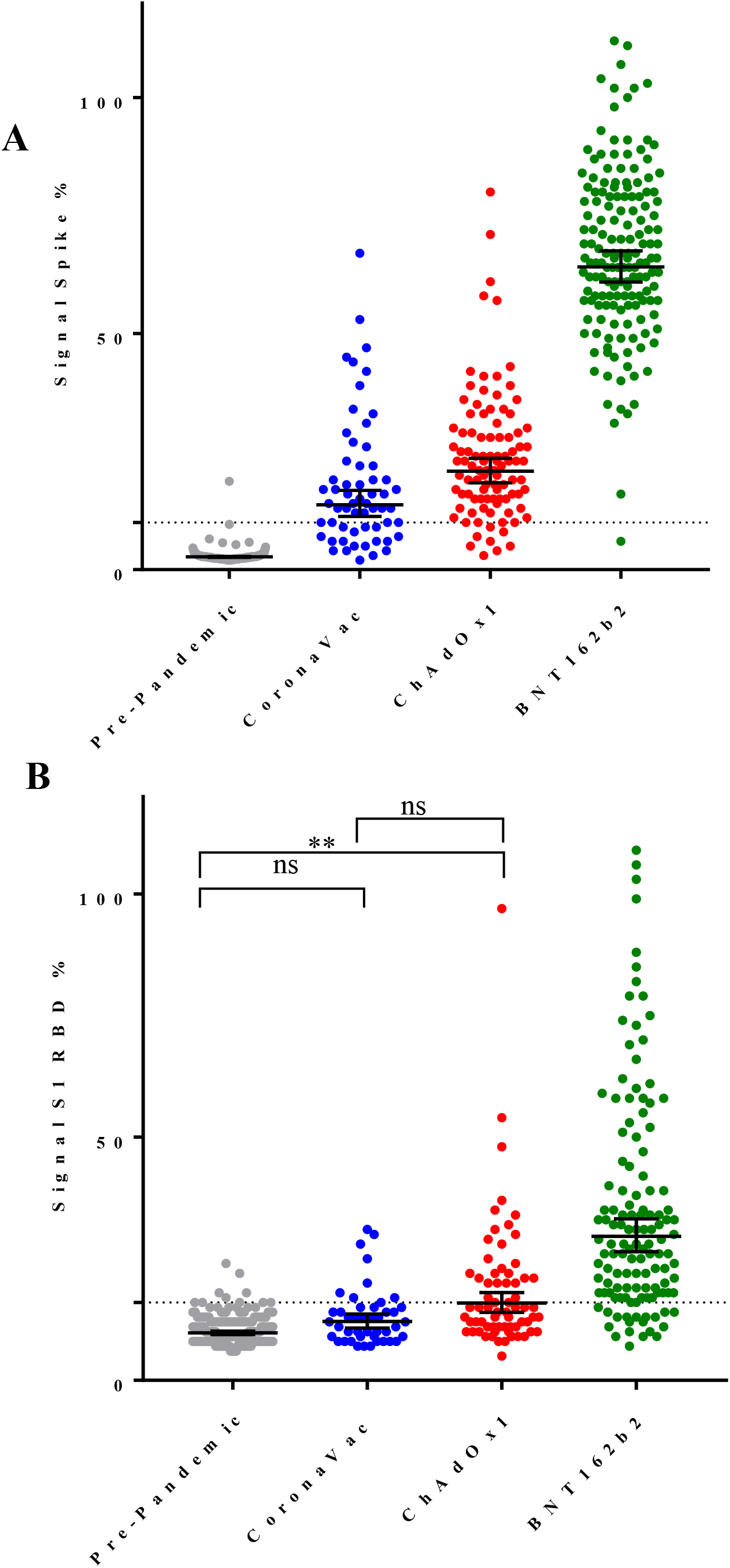
The IgG response to Spike and S1 RBD accordingly to the vaccine type. IgG levels reactive for Spike (A) and S1 RBD (B) in participants negative for COVID-19, with 18 to 40 years of age which completed primary vaccination (2^nd^ dose) between 10 to 90 days. The dashed line indicates the seropositive assay cutoff. Pre-pandemic samples were plotted as naïve controls. The geometric mean and 95% CI is represented by the bars. One-Way ANOVA analysis were performed (**** p<0.0001), (**p<0.01), (ns, not significant).

**Fig. S6.**
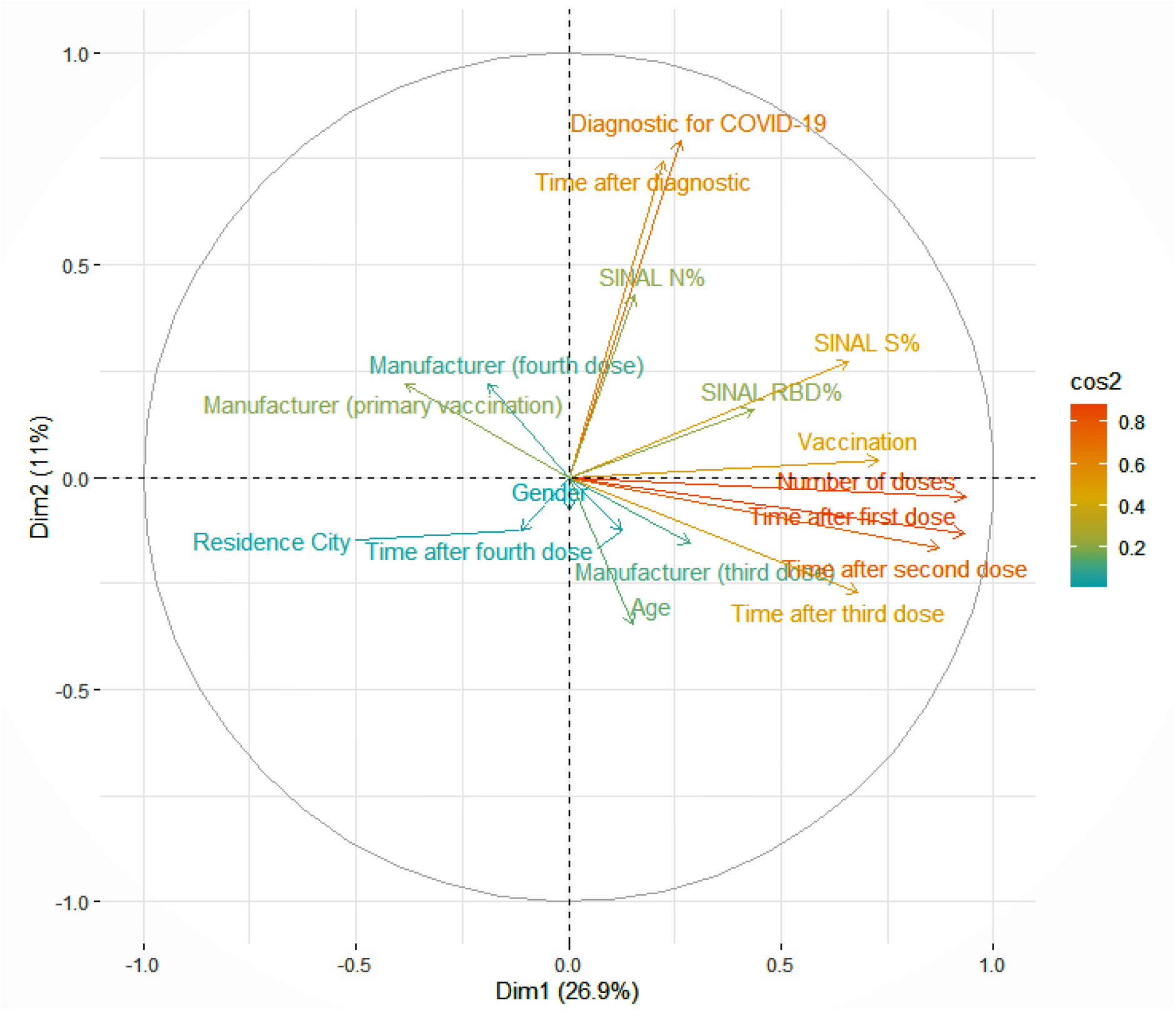
Principal Component Analysis (PCA). The table of variables was subject to PCA analysis. The influence of each variable in the variance of the dataset along PC1 and PC2 are indicated as vectors. Positively correlated vector variables are grouped together. The quality of representation of each variable for the variance of the dataset is indicated by the color gradient (strong to weak, red to blue)

